# National and state wise estimate of time varying reproduction number for COVID-19 in India during the nationwide lockdown

**DOI:** 10.1101/2020.05.01.20087197

**Authors:** Padmanaban Venkatesan

## Abstract

To understand the effect of nationwide lockdown on transmissibilty of SARS-CoV-2 in India, time varying reproduction number during the first weeks of April, 2020 was estimated. The time varying reproduction number was estimated using EpiEstim package in R programming language. The reproduction number has come down significantly during the lockdown period both at national level and in most states but it wasn’t reduced to less than 1. This calls for urgent need for more effective control measures in addition to lockdown to stop the epidemic spread of the virus.

## Introduction

Central government of India implemented a nationwide lockdown from midnight of March 24, 2020 as a response to growing threat of COVID-19 pandemic caused by a novel corona virus (SARS-CoV-2 virus) (1). Despite implementing one of the strictest lockdowns in the world, the reported number of COVID-19 cases in India continue to increase even after more than 5 weeks of lockdown (2). This has led to questions on the effectiveness of the lockdown in controlling the spread of the virus (3). Further, there are inter-state differences in spread of the virus with some states reporting lower number of cases than before and others reporting surge in incidence of COVID-19 cases.

In this study, to understand the transmissibilty of the virus during the lockdown period in India and in individual states, we measured time varying reproduction number for the first 4 weeks of April, 2020.

## Method

We estimated time varying reproduction number for India and individual states by implementing the package “EpiEStim” in R programming language (4). The method used in the package was described by Cori. et.al. (5). Total and state wise incidence of COVID-19 cases during 1^st^ April to 28^th^ April was retrieved from the crowdsourced web platform covid19india.org (2). Mean genertion time of 5.2 days and SD of 2.8 days as reported from China was used to estimate the reproduction number (6).

## Results

Table 1 shows the estimated reproduction number for the first(2^nd^ to 8^th^) and fourth week(22^nd^ to 28^th^) of April, 2020 for each state and India. Overall for India, the reproduction number came down from 2.39 during first week to 1.19 during last week of April.Due to the time between infection and symtopm onset and delay between symptomp onset and testing and reporting of results, the reproduction number during first week of April might reflect the reproduction number during pre-lockdown days. Also the reproduction number estimate for initial weeks might be a more of an underestimate than later weeks because of comparatively much lower testing rate during the earlier days. Eventhough the reprodution number has decreased significantly which would result in ‘flattening’ of the epidemic curve, it is still not less than 1 to effectively stop the curve. Among the states, Bihar and Jharkhand continue to have high reproduction number. Major states such as Maharashtra, Madhya Pradesh, Delhi and West Bengal have higher reproduction number than national average. Karnataka, Rajasthan, UP and Telangana show less than 1 reproduction number but it could be result of poor testing rate for SARS CoV-2 in some of these states. In particular, Telangana have reduced the number of tests done per day even when number of cases reported was increasing during the previous days (7,8). This resulted in a sudden decrease in number of new reported cases and thus a very low reproduction number of 0.35 during the last week of April.

**Table 1:** Reprouduction number of SARS-CoV-2 during first and fourth week of April, 2020

| States | 2<sup>nd</sup> to 8<sup>th</sup> April |  |  | 22<sup>nd</sup> to 28<sup>th</sup> April |  |  |
| --- | --- | --- | --- | --- | --- | --- |
|  | Median R | 95% Confidence interval |  | Median R | 95% Confidence interval |  |
|  |  | LL | UL |  | LL | UL |
| Andhra Pradesh (AP) | 1.78 | 1.57 | 2.02 | 1.45 | 1.33 | 1.58 |
| Bihar (BR) | 1.90 | 1.11 | 3.01 | 2.18 | 1.92 | 2.47 |
| Chhattisgarh (CT) | 2.46 | 0.35 | 8.15 | 0.93 | 0.21 | 2.50 |
| Delhi (DL) | 2.16 | 1.98 | 2.35 | 1.52 | 1.44 | 1.61 |
| Gujarat (GJ) | 3.14 | 2.56 | 3.79 | 1.03 | 0.98 | 1.09 |
| Haryana (HR) | 4.40 | 3.68 | 5.22 | 0.99 | 0.75 | 1.28 |
| Jammu and Kashmir (JK) | 3.41 | 2.77 | 4.13 | 1.43 | 1.23 | 1.64 |
| Jharkhand (JH) | 2.63 | 0.78 | 6.29 | 2.18 | 1.67 | 2.78 |
| Karnataka (KA) | 2.22 | 1.74 | 2.77 | 0.81 | 0.67 | 0.98 |
| Kerala (KL) | 1.17 | 0.90 | 1.49 | 1.01 | 0.76 | 1.31 |
| Madhya Pradesh (MP) | 2.75 | 2.41 | 3.10 | 1.31 | 1.22 | 1.40 |
| Maharashtra (MH) | 3.08 | 2.87 | 3.30 | 1.29 | 1.25 | 1.33 |
| Orissa (OR) | 2.25 | 1.61 | 3.05 | 1.35 | 0.97 | 1.82 |
| Punjab (PB) | 3.62 | 2.79 | 4.61 | 1.15 | 0.93 | 1.40 |
| Rajasthan (RJ) | 2.67 | 2.36 | 3.01 | 0.84 | 0.77 | 0.91 |
| Tamil Nadu (TN) | 1.79 | 1.64 | 1.96 | 1.11 | 1.01 | 1.22 |
| Telangana (TG) | 2.38 | 2.13 | 2.65 | 0.35 | 0.28 | 0.43 |
| Uttar Pradesh (UP) | 2.57 | 2.26 | 2.91 | 1.00 | 0.93 | 1.08 |
| West Bengal (WB) | 3.02 | 2.33 | 3.83 | 1.29 | 1.15 | 1.44 |
| India | 2.39 | 2.32 | 2.47 | 1.19 | 1.17 | 1.21 |

Figure 1 to 6 show the change in weekly reproduction number of major states and India. The reproduction number against a particular date is for a week with the last day of the week ending on the particular date.

**Figure 1:**
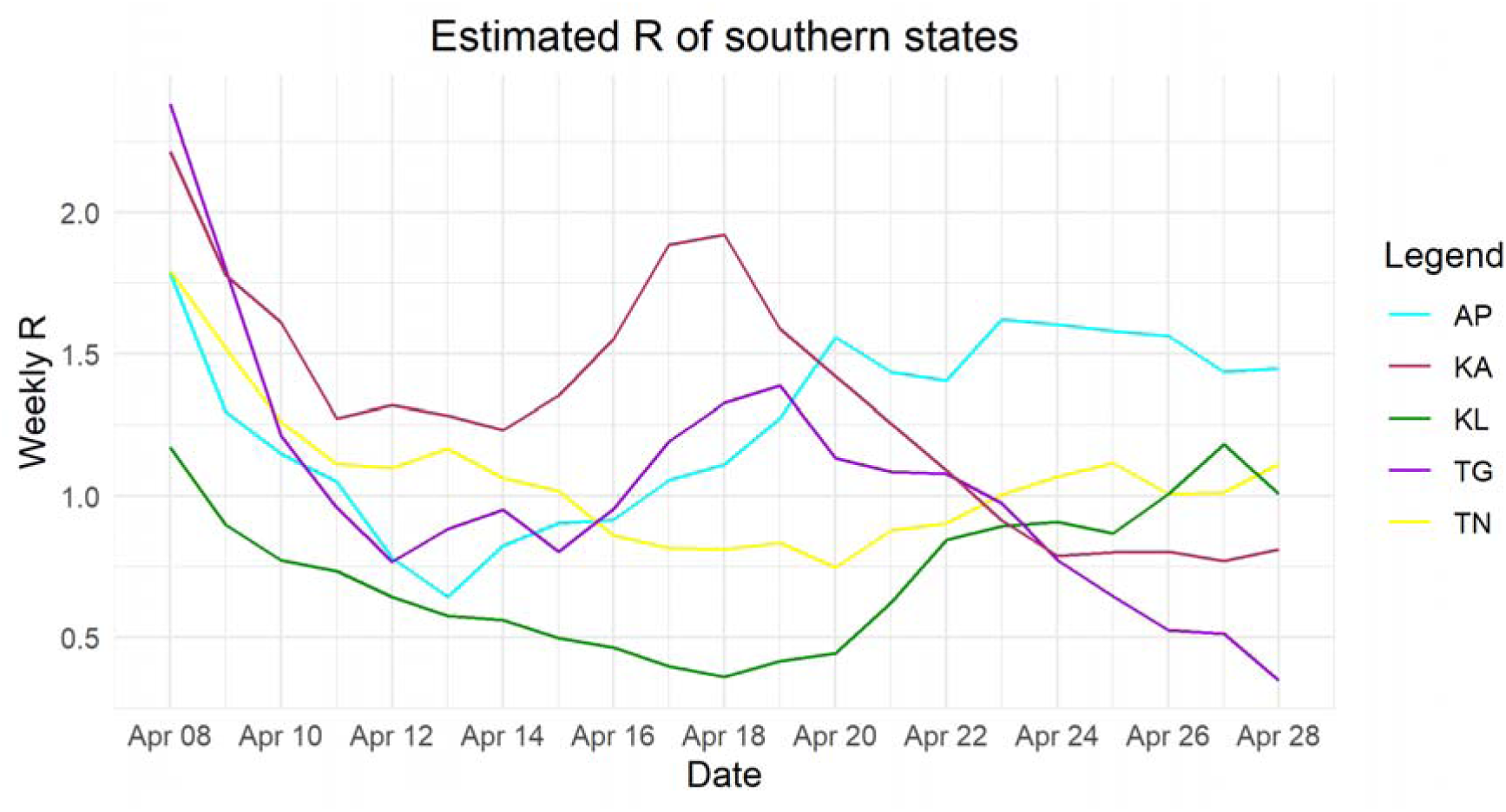
Estimated weekly reproduction number of southern states.

**Figure 2:**
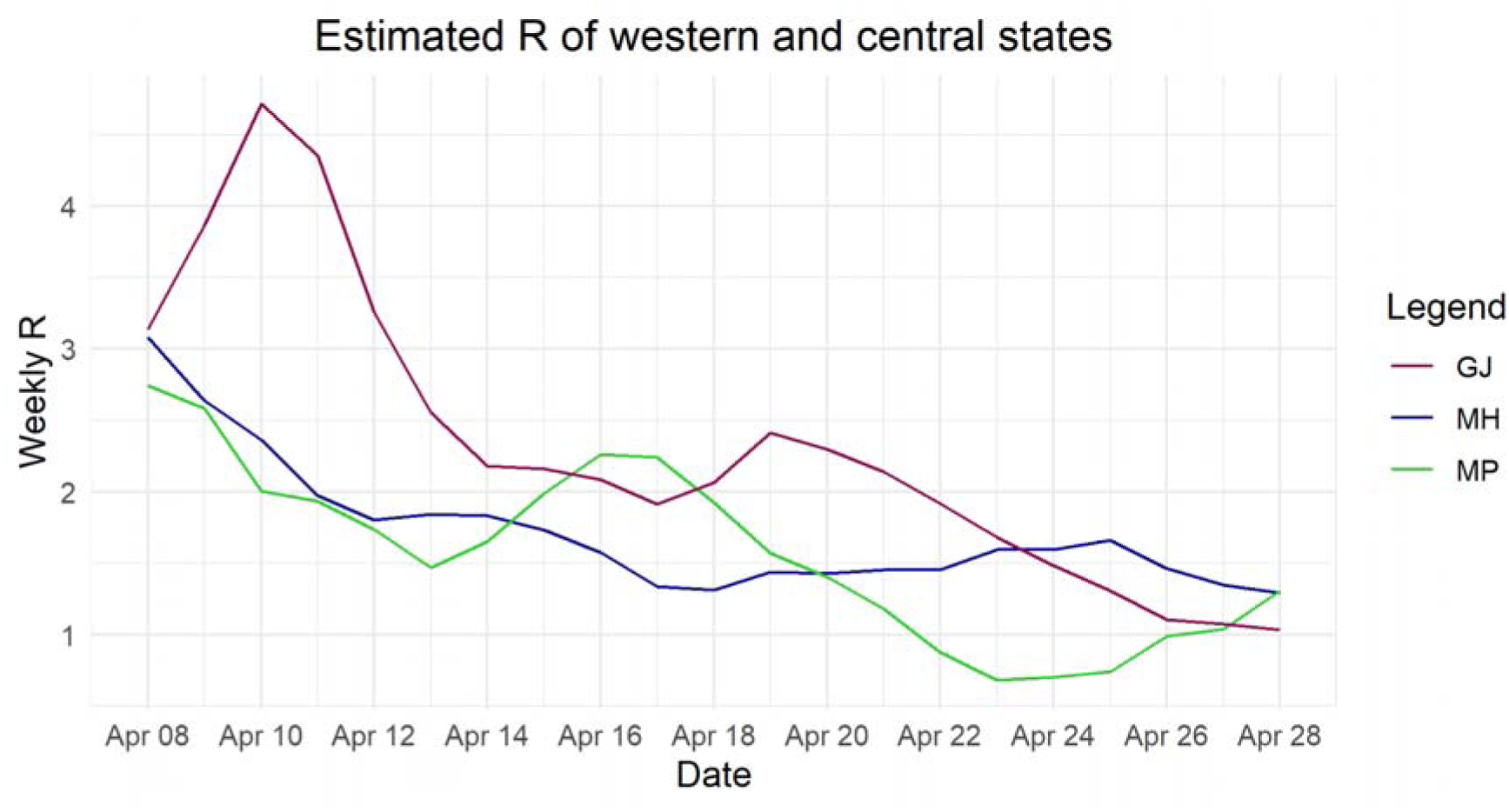
Estimated weekly reproduction number of western and central states.

**Figure 3:**
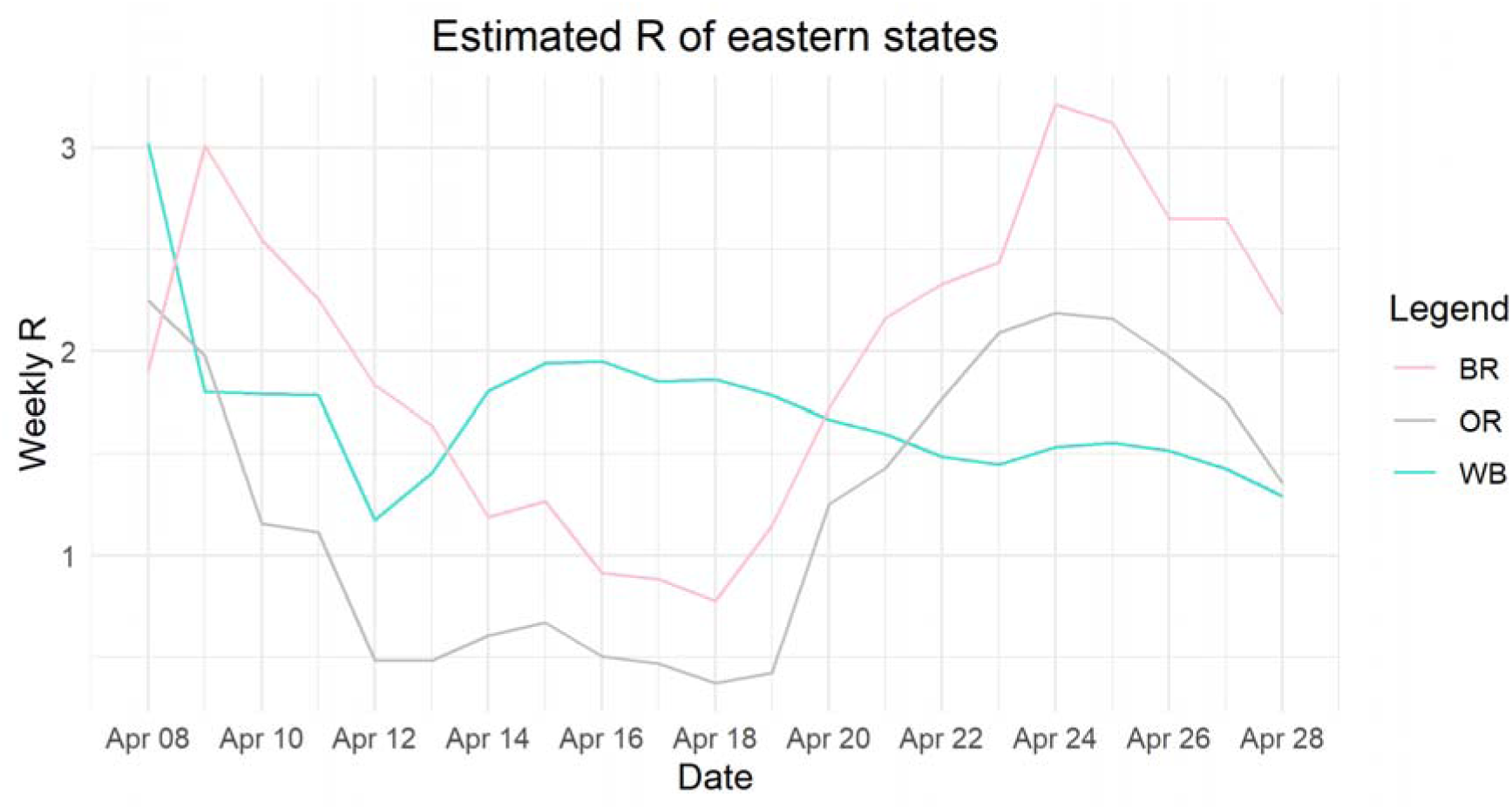
Estimated weekly reproduction number of eastern states.

**Figure 4:**
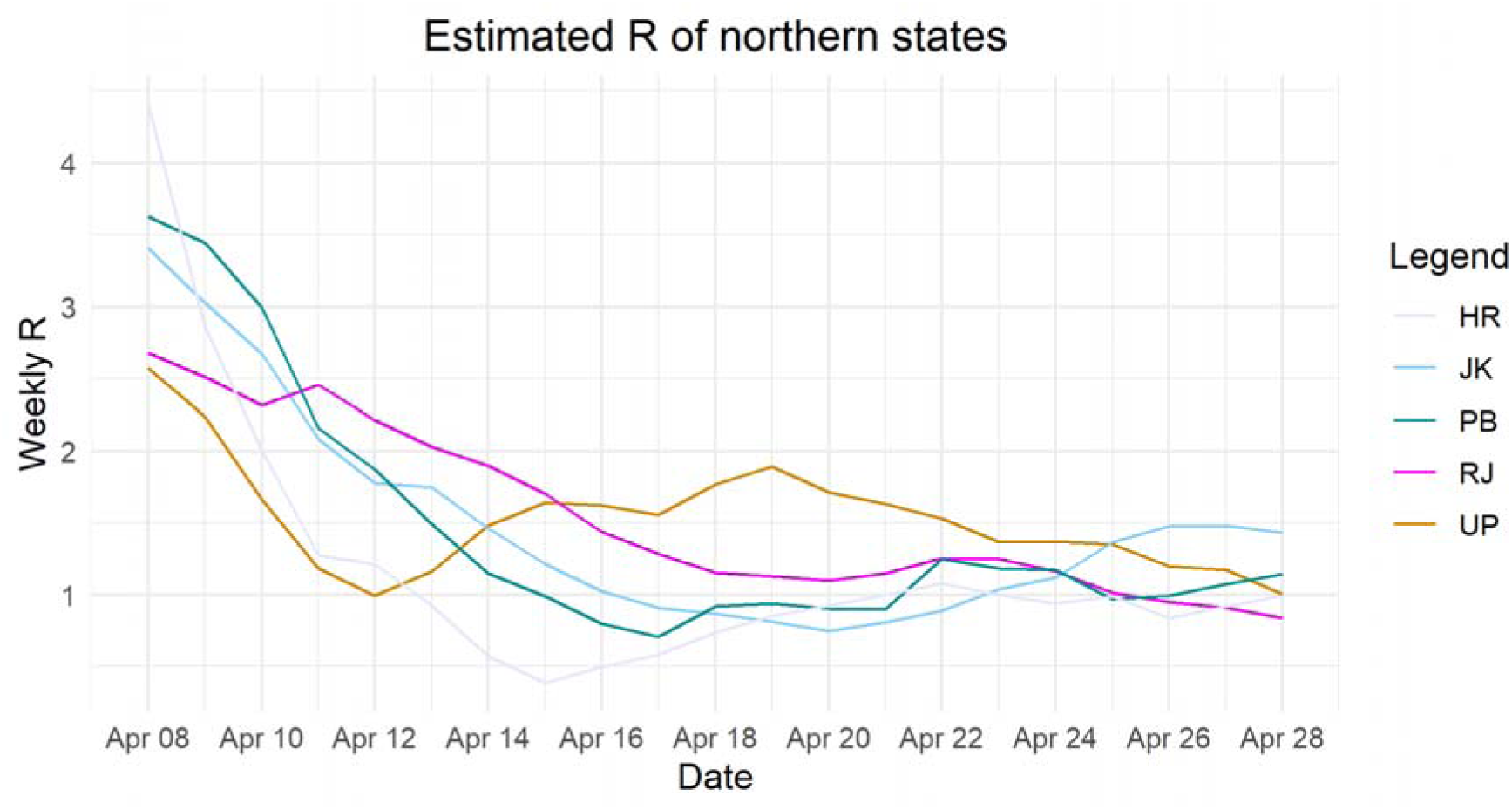
Estimated weekly reproduction number of northern states.

**Figure 5:**
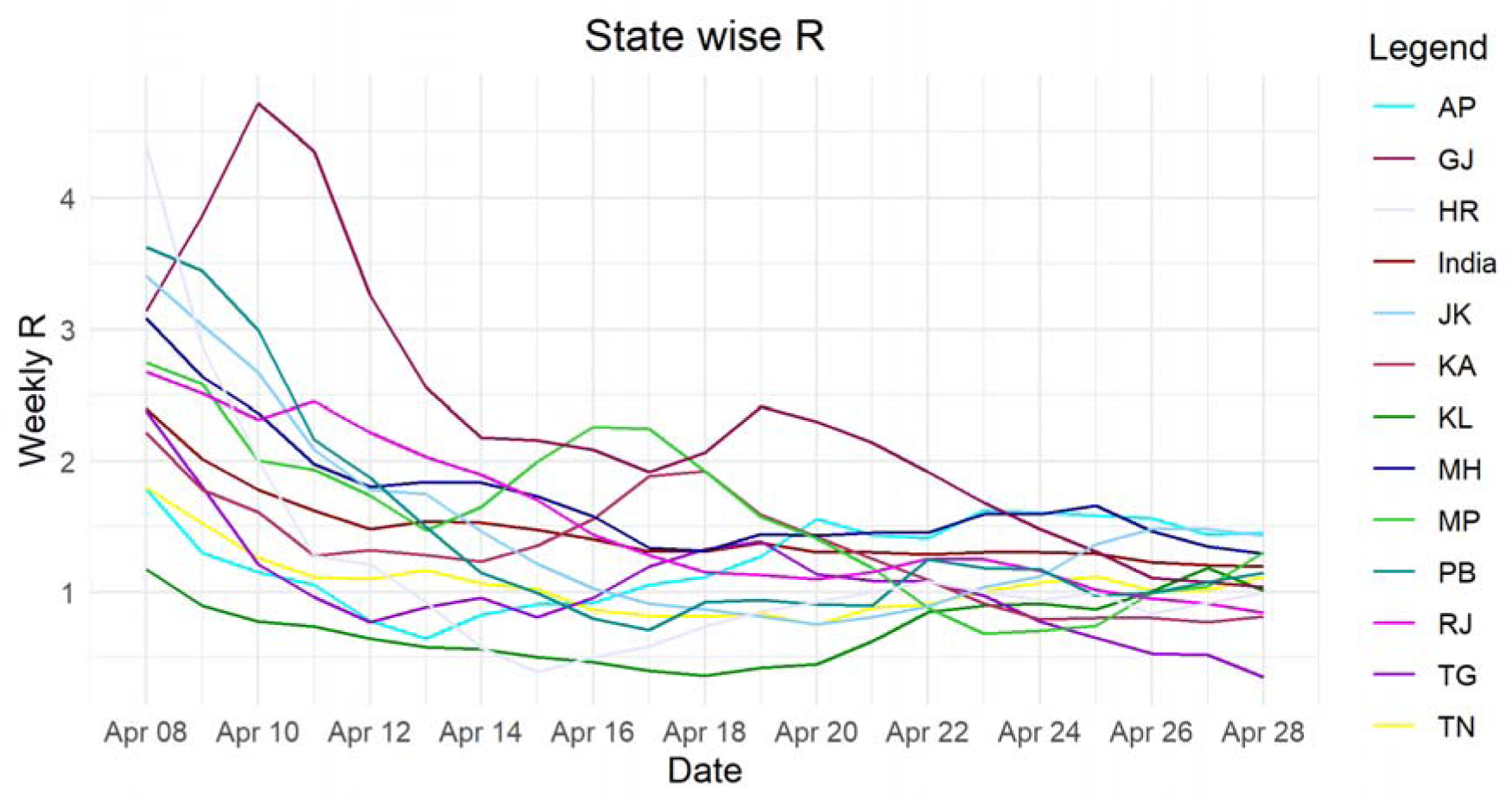
Estimated weekly reproduction number of major states.

**Figure 6:**
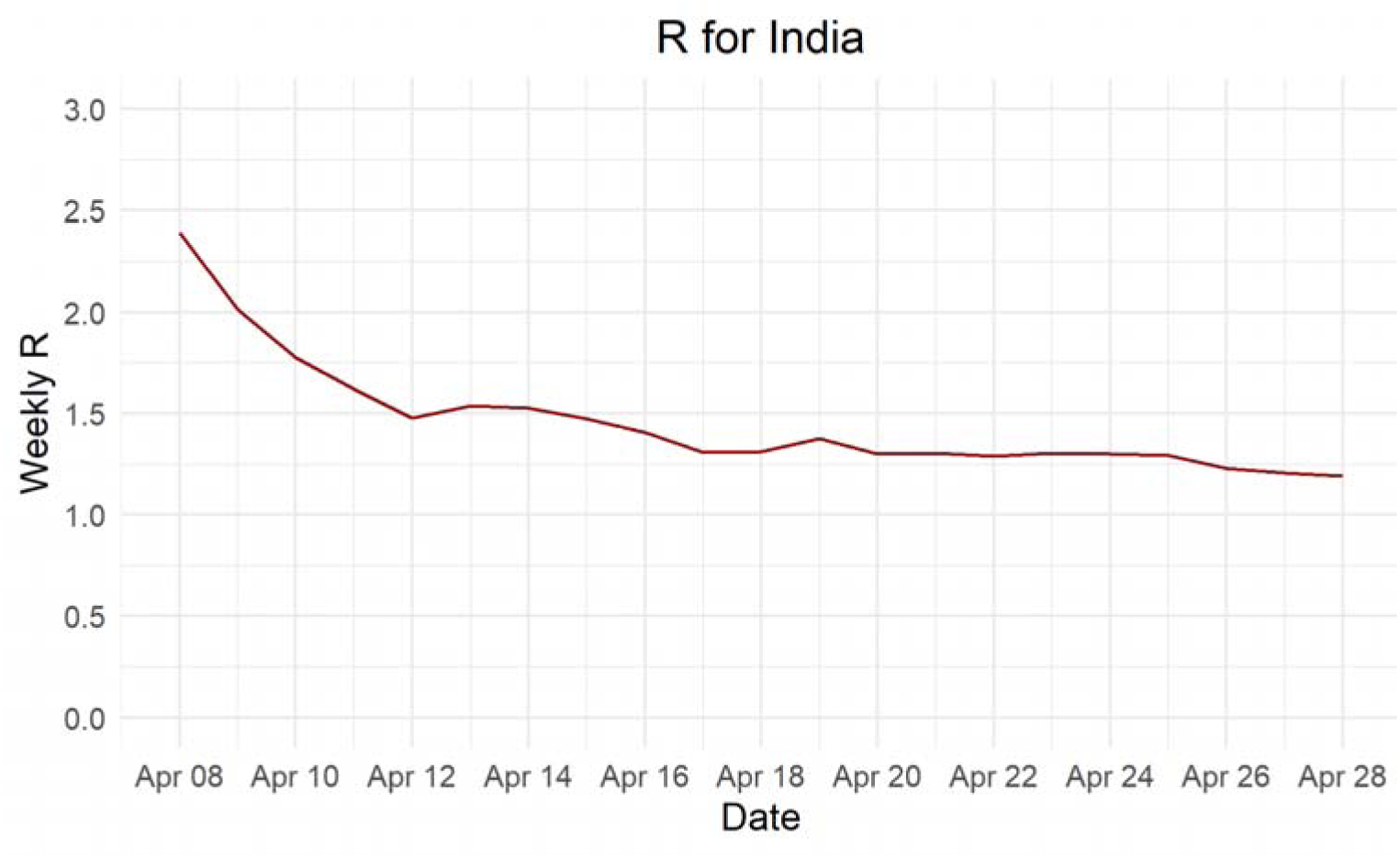
Estimated weekly reproduction number of India.

## Conclusion

There is singificant decrease in the reproduction number during the lockdown but it is not reduced to less than 1 to stop the epidemic. This underlies the fact that lockdown alone is not enough to stop the epidemic in India and other measures such as extensive testing, contact tracing and isolation of COVID-19 positve subjects is essential. Limitations of the estimation include poor testing rate and variations in testing rate between states. Since enough data is not publicly available to estimate generation time for COVID-19, the generation time reported by a study from China was used and this could limit the accuracy of the estimates for the reprodcution number.

## Data Availability

Data used for the study is publicly available

## Notes

### Competing Interest Statement

The authors have declared no competing interest.

### Funding Statement

No funding was received for this study

